# Heterozygous *PRKN* mutations are common but do not increase the risk of Parkinson’s disease

**DOI:** 10.1101/2021.08.11.21261928

**Authors:** William Zhu, Xiaoping Huang, Esther Yoon, Sara Bandres-Ciga, Cornelis Blauwendraat, Joshua H. Cade, Beverly P. Wu, Victoria H. Williams, Alice B. Schindler, Janet Brooks, J Raphael Gibbs, Dena G Hernandez, Debra Ehrlich, Andrew Singleton, Derek Narendra

**Affiliations:** Inherited Disorders Unit, Neurogenetics Branch, Division of Intramural Research, National Institute of Neurological Disorders and Stroke, National Institutes of Health, Bethesda, MD, USA; Parkinson’s Disease Clinic, Office of the Clinical Director, National Institute of Neurological, Disorders and Stroke, National Institutes of Health, Bethesda, MD, USA; Molecular Genetics Section, Laboratory of Neurogenetics, National Institute on Aging, National Institutes of Health, Bethesda, MD, USA; National Institute of Neurological Disorders and Stroke, Neurogenetics Branch, National Institutes of Health, Bethesda, MD, USA; Integrative Neurogenomics Unit, Laboratory of Neurogenetics, National Institute on Aging, National Institutes of Health, Bethesda, MD, USA; Center for Alzheimer’s and Related Dementias, National Institutes of Health, Bethesda, MD

**Keywords:** PARK2, Parkin, Mitophagy, Early Onset Parkinson’s Disease, Young Onset Parkinson’s Disease

## Abstract

*PRKN* mutations are the most common recessive cause of Parkinson’s disease (PD) and are a promising target for gene and cell replacement therapies. Identification of biallelic *PRKN* patients (PRKN-PD) at the population scale, however, remains a challenge, as roughly half are copy number variants (CNVs) and many single nucleotide polymorphisms (SNPs) are of unclear significance. Additionally, the true prevalence and disease risk associated with heterozygous *PRKN* mutations is unclear, as a comprehensive assessment of *PRKN* SNPs and CNVs has not been performed at a population scale. To address these challenges, we evaluated *PRKN* mutations in 2 cohorts analyzed with both a genotyping array and exome or genome sequencing: the NIH PD cohort, a deeply phenotyped cohort of PD patients, and the UK Biobank, a population scale cohort with nearly half a million participants. Genotyping array identified the majority of *PRKN* mutations and at least 1 mutation in most biallelic *PRKN* mutation carriers in both cohorts. Additionally, in the NIH-PD cohort, functional assays of patient fibroblasts resolved variants of unclear significance in biallelic carriers and ruled out cryptic loss of function variants in monoallelic carriers. In the UK Biobank, we identified 2,692 *PRKN* CNVs from genotyping array data from nearly half a million participants (the largest collection to date). Deletions or duplications involving exons 2 accounted for roughly half of all CNVs and the vast majority (88%) involved exons 2, 3, or 4. Combining estimates from whole exome sequencing (from ∼200,000 participants) and genotyping array data, we found a pathogenic *PRKN* mutation in 1.8% of participants and 2 mutations in ∼1/7,800 participants. Those with 1 *PRKN* pathogenic variant were as likely as non-carriers to have PD (OR = 0.91, CI= 0.58 – 1.38, p-value = 0.76) or a parent with PD (OR = 1.12, CI = 0.94 – 1.31, p-value = 0.19). Together our results demonstrate that heterozygous pathogenic *PRKN* mutations are common in the population but do not increase the risk of PD. Additionally, they suggest a cost-effective framework to screen for biallelic *PRKN* patients at the population scale for targeted studies.

## INTRODUCTION

Parkinson’s disease (PD) is the second most common neurodegenerative disorder with a prevalence of about 0.5% in individuals ≥ 45 years of age ^1^. 5 -10% of cases are caused by mutation(s) in a single gene ^2^. Mutations in *PRKN* are the most common recessive form of PD (*PRKN*-PD), and *PRKN*-PD, in particular, is a promising target for gene and cell replacement therapies ^3–5^. Patients with *PRKN*-PD typically have disease onset before the age of 40 ^6^.

For gene-targeted trials, the target gene needs to be identified in a large group of individuals, ideally early in the disease course. Cost-effective genotyping platforms, notably genotyping arrays, have facilitated screening for mutation carriers for clinical trials, sometimes through partnership with consumer-based genotyping companies ^7^. Genotyping arrays are particularly effective for mutations such as *APOE* E4 or *LRRK2* p.G2019S in which the locus can be genotyped from 1 or a few single nucleotide polymorphisms (SNPs).

Identification of individuals with recessive disorders like *PRKN*-PD by genotyping array, however, presents additional challenges. Recessive disorders are typically caused by loss of function (LOF) variants that may be scattered throughout the gene, 2 pathogenic variants must be identified to establish causality, and both SNPs and copy number variants (CNVs) may lead to loss of function, necessitating analytic methods that can detect both. In the face of these challenges, how well genotyping arrays capture *PRKN* mutations in PD cohorts and the general population is largely unknown.

The challenges of identifying *PRKN* mutations at population scale has additional implications. Because large scale studies do not typically capture all *PRKN* SNPs and/or all CNVs, the true prevalence of *PRKN* mutations in the general population is not known with estimates ranging from 0.17% to 3.7% ^8^. Additionally, some studies have suggested that single *PRKN* mutation carriers have an increased risk of developing idiopathic PD (iPD) ^9^. These case-control studies, however, may be confounded by cases with a missed second *PRKN* mutation. Notably other studies have failed to find this association and a recent meta-analysis suggested that the association may depend on a missed mutation in biallelic variant carriers ^10^. Improving genotyping of *PRKN* mutations at the population scale may help clarify the prevalence of *PRKN* in the general population and the risk of iPD conferred by a single *PRKN* mutation.

Finally, in the context of a *PRKN* gene targeted trial, it may be important to determine definitively whether an individual patient is carrying two pathogenic *PRKN* mutations. This can be challenging if a rare *PRKN* variant is found without proven disease segregation. Similarly, it can be challenging to know whether a cryptic variant may be missed in a PD patient with only 1 detected pathogenic *PRKN* variant. Cellular assays to assess *PRKN* function in patient cells may be helpful in these cases. *PRKN* functions can be monitored in cells, such as its ubiquitination of substrates like mitofusin-1 (MFN1), its production of phospho-ubiquitin, or its promotion of mitophagy ^11–19^. Notably, MFN1 ubiquitination has been used to evaluate PARKIN function in patient fibroblasts in small numbers of patients or single families ^16,20^. How well these assays distinguish unrelated patients with 1 or 2 *PRKN* mutations in patient fibroblasts with endogenous levels of *PRKN*, however, has not been assessed in a large series.

To help address these challenges, we first validated genotyping arrays for identifying *PRKN* mutations in a large PD cohort, using whole genome sequencing (WGS) as the gold standard. We then recruited identified patients with 1 or 2 *PRKN* mutations to the PD clinic and assessed PARKIN function in their fibroblasts. Finally, we assessed *PRKN* mutations in the UK Biobank, a large population study with genotyping array data available from nearly half a million participants and whole exome sequencing (WES) data from ∼200,000 participants ^21^. Together our results show that there is a high prevalence of heterozygous pathogenic *PRKN* mutations in the general population and that they do not increase PD risk. Additionally, our findings suggest a screening framework to identify biallelic *PRKN* patients for targeted studies with high confidence.

## MATERIALS AND METHODS

### NIH-PD Cohort

Study participants were recruited to the Parkinson’s Disease Clinic of the National Institute of Neurological Disorders and Stroke (NINDS) and the National Institutes of Health Clinical Center. All participants gave written informed consent according to the Declaration of Helsinki to protocols approved by the Institutional Review Board of NINDS before undergoing research procedures.

### Functional analysis of patient fibroblasts

Patient fibroblast lines were established from 3-mm punch biopsies taken from the forearm. Cell lines were assayed at passage 11 or before. For analysis of MFN1 and pS65-Ub, cells treated with DMSO or valinomycin 10 mM overnight (>16 hrs) were washed in PBS and pelleted. Cell pellets were lysed in RIPA buffer on ice for 30 minutes and then cleared by centrifugation at 21,130 g. Protein concentration was determined using the BCA assay. 20 mg of protein was loaded for each sample. Samples were separated on 7.5% Criterion TGX precast midi protein gel (Biorad) and proteins were transferred to nitrocellulose membranes. Membranes were blotted with MFN1 using an antibody from Proteintech (cat no., 13798-1-AP). Some of the same lysates were blotted for pS65-ubiquitin (Sigma, cat no. ABS1513). Indicated patient fibroblast lines were transduced with mt-Keima and analyzed by flow cytometry as previously reported ^22^. For examination of *PRKN* exon 1 – exon 2 splicing, RNA was isolated from the indicated fibroblast lines using the Direct-zol RNA miniprep kit (RPI research products, cat no. ZR2051) and reverse transcribed using a kit from Thermofisher (cat no., 4368814). The *PRKN* exon 1 – 3 product was amplified with MyTaq DNA polymerase (Bioline) from 60 ng of cDNA using the following primers: 5’-aggatttaacccaggagagc -3’ and 5’-aatgctctgctgatccaggt -3’. b-actin was amplified using primers from Thermofisher (cat no., cat# Hs01060665_g1).

### Genotyping NIH-PD Cohort

Genotyping of the NIH-PD cohort was performed using a genome-wide coverage genotyping array (NeuroX or Neuro Consortium Array, Illumina, Inc., San Diego, CA) and/or WGS. The NeuroX array is based on the Illumina HumanExome array v1.1 and the NeuroChip array is based on the Infinium HumanCore-24 c1.0 array ^23,24^. Both have additional custom content covering neurodegenerative disease-related variants. To identify SNPs from the genotyping array, Illumina GenomeStudio (v.2.0) was used cluster genotypes. Quality-Control measured included limiting to samples with call rates of >95%, excluding samples with excess heterogeneity (F statistic ± 0.25), and excluding samples whose genotyped sex did not match the sample demographics. CNVs were identified by manual inspection of the B Allele Frequency and Log R Ratio for the *PRKN* gene region (Chr6: 161770811 -163140694, hg19), using the gglot2 visualization package for R (https://www.r-project.org/), as described previously ^25^.

For WGS, 1 mg of total genomic DNA was sheared to a target size of 450 base pairs (bp) by ultrasonication. The library was prepped with the TruSeq DNA PCR-Free High Throughput Library Prep Kit and IDT for Illumina TruSeq DNA UD Indexes (96 Indexes, 96 Samples). Samples were sequenced on an Illumina HiSeq X System using 150 bp paired end reads. Called SNPs were annotated using ANNOVAR ^26^. CNVs in the *PRKN* region were detected using the Manta structural variant caller (Illumina) ^27^. For cases with available DNA, detected *PRKN* CNVs were verified by multiplex ligation dependent probe amplification with SALSA MLPA Probemixes P051 and P052 (MRC Holland).

### UK Biobank data

To identify CNVs in the UK Biobank data ^28^, the UK Biobank B allele frequency (BAF) and Log R Ratio data were downloaded from UK Biobank (v2), containing data from 488,377 participants. BAF and Log R Ratio data were extracted for the *PRKN* gene region (Chr6: 161770811 -163140694, hg19). Potential CNVs were detected using three algorithms. The first, *PRKN* del finder 1, averaged the log_2_ ratio intensity for SNPs within or immediately flanking each exon of *PRKN*. It flagged an exon as possibly deleted if the exon average was 1.5s less than the average of all the exons or possibly duplicated if it was greater than 2s the average of all the exons. The second algorithm, *PRKN* dup finder, divided the *PRKN* locus into 12 regions flanking each *PRKN* exon and then counted the number of SNPs with BAF in the ranges of 0.125 -0.375 and 0.625 -0.875. Samples were flagged if ≥ 2 positive SNPs were identified in any region. Finally, the third algorithm, *PRKN* del finder 2, counted the number of SNPs with low Log R Ratio intensities in these same regions flanking each exon. Samples were flagged if there were ≥ 3 positive SNPs in any region. All flagged samples were then assessed visually for CNVs by plotting the BAF and Log R Ratio values with the gglot2 visualization package for R (https://www.r-project.org/). The following phenotypic data was obtained from the UK Biobank: ICD10 codes (field code: 41270), PD (field code: 131023), parkinsonism (field code: 42031), illnesses of father and mother (field codes: 2017 and 20110), genetic ethnic grouping (field code: 22006), year of birth (field code: 34), and age of recruitment (field code: 21022). UK Biobank exome sequencing data (FE dataset, field code: 23156) were downloaded from the UK Biobank. Variants were annotated using ANNOVAR ^26^. Pathogenicity of variants was determined using their ClinVar annotation (https://www.ncbi.nlm.nih.gov/clinvar/). Likely pathogenic variants were grouped with pathogenic and likely benign variants were groups with benign. We inspected the evidence for variants annotated as “conflicting interpretations of pathogenicity” and categorized as pathogenic if most reports listed the variant as pathogenic or likely pathogenic and benign if most reports listed it as benign or likely benign. The primary association test for PD risk due to *PRKN* variants was testing whether having 1 pathogenic *PRKN* variant identified by WES or genotyping array increases risk of PD relative to having 0 pathogenic *PRKN* variants. Additional tests were performed to assess sensitivity of the analysis including testing whether 1 *PRKN* variant increases the risk of having a parent with PD, testing different classes of variants separately, and testing detected variants in the larger sample with just genotyping array data. As these secondary sensitivity analyses were considered exploratory, reported p-values were not adjusted for multiple comparisons. No assumptions about Hardy-Weinberg equilibrium were made. No methods were used to infer genotypes or haplotypes. No specific methods were used to address population stratification. No method was used to address relatedness among participants.

### Statistical analyses

In analyses of patient fibroblasts, statistical significance was determined using one-way ANOVA followed by Tukey’s multiple comparisons test implemented in Prism 9 (GraphPad). Odds ratio and p-value for contingency tables was calculated in R using Fischer’s Exact Test (https://www.r-project.org/). For calculating the frequency and odds ratio for each mutation, samples with missing data were omitted.

### Data and code availability

UK Biobank data is publicly available upon application at the UK Biobank website (https://www.ukbiobank.ac.uk/). gnomAD v2.1.1 summary statistics are available from (https://gnomad.broadinstitute.org/). MDS database summary statistics are available from (http://www.mdsgene.org/). Code used for analysis is available on our GitHub repository (https://github.com/NarendraLab/Parkin/).

## RESULTS

### Identification of biallelic PRKN patients in NIH PD cohort by WGS

*PRKN* mutations were assessed by WGS in a discovery cohort of 742 patients seen consecutively at the NIH from 2006 – 2019 and for whom DNA was available (hereafter, the NIH PD cohort) (Figure 1A). A total of 18 known pathogenic SNPs and 13 exon-spanning deletions in *PRKN* were identified (Figure 1B (black font) and Table 1). Multiplex ligation-dependent probe amplification verified the deletions in 12/12 samples that were available for testing.

**Figure 1.**
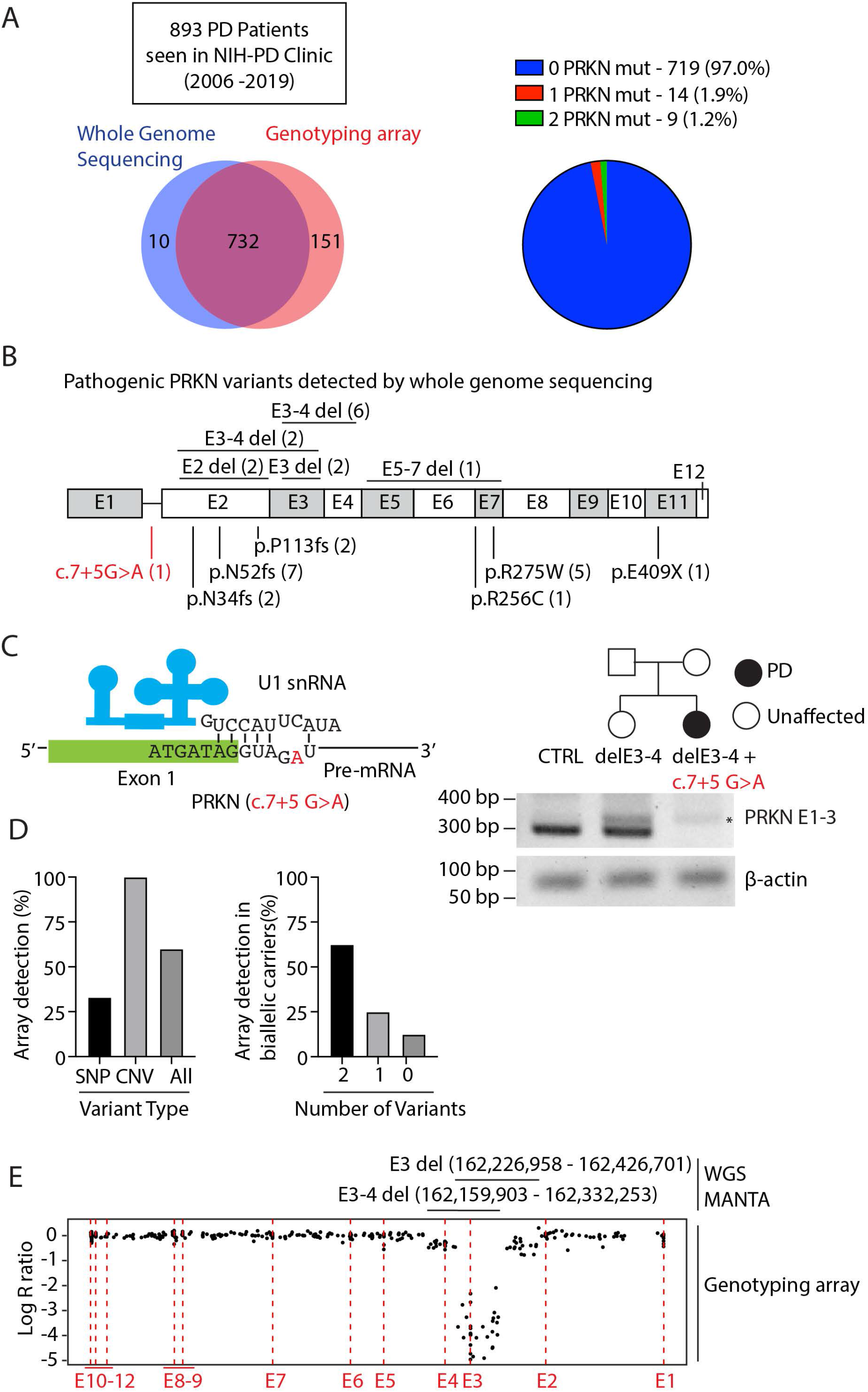
*PRKN* variants in the NIH PD cohort. (A) Consecutive PD cases seen in the NIH-PD clinic between years 2006 and 2019 were genotyped by genotyping array, whole genome sequencing, or both as depicted in Venn diagram (left). Proportion of *PRKN* variants detected in the NIH-PD cohort by whole genome sequencing is shown in the pie chart (right). (B) The position of pathogenic *PRKN* variants (black) and a novel VUS (red) detected by whole genome sequencing are shown in schematic of the *PRKN* gene. CNVs are shown on the top and SNPs on the bottom. (C) Schematic of the exon 1 – intron 1 junction is shown with the position of the intronic substitution c.7+5 G>A in red (left). This is predicted to disrupt a base pairing with the U1 snRNA of the spliceosome. Pedigree of family with this mutation appears on upper right. RT-PCR of RNA isolated from fibroblast cell lines from a control (CTRL), the unaffected sister of the proband with a single E3-4 del, and the proband with an E3-4 del and c.7+5 G>A variants is depicted on the lower right. The c.7+5 G>A mutation disrupted the PCR product from primers in exons 1 and 3. An aberrant product (^*^) was seen in both E3-4 del carriers. b-actin served as a loading control. (D) Graph on left represents the proportion of *PRKN* variants (either single nucleotide polymorphisms (SNPs) or copy number variants (CNVs)) detected by genotyping array. Graph on right represents the number of *PRKN* variants detected by genotyping array in biallelic *PRKN* patients. (E) Correspondence between CNV calls by whole genome sequencing (WGS) and by genotyping array is shown for a representative case with compound heterozygous *PRKN* deletions. The x-axis represents the relative genomic position. The position of the exons is indicated by the red dotted lines. Log R ratio is shown on the y-axis. A value of -0.5 corresponds to a single deletion and a lower value is expected for two deletions. The relative position of the deletions called from WGS is shown above. Log R ratio is around -0.5 for probes in the non-overlapping regions of the deletions and < -2 for probes in the overlapping region.

**Table 1:**
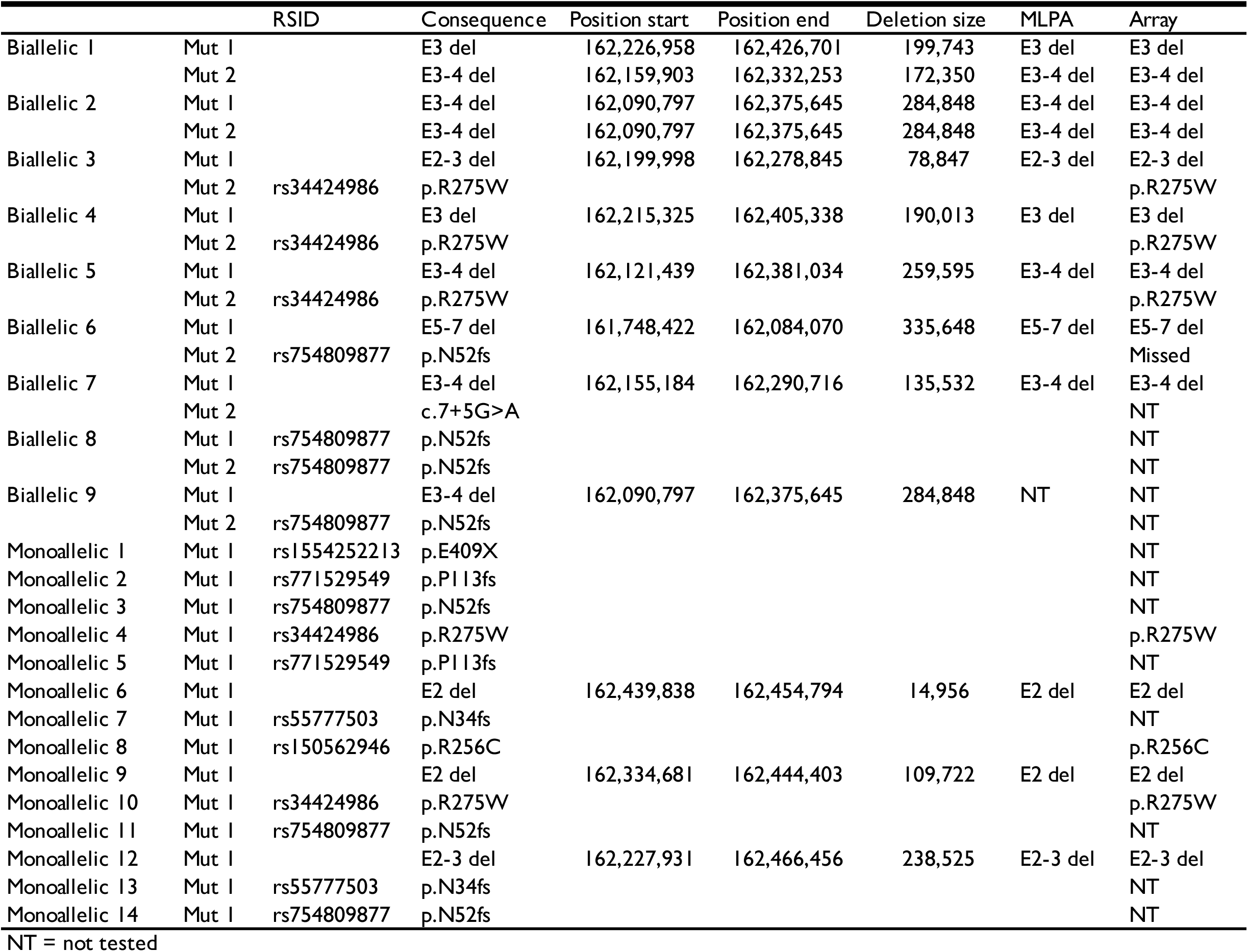
Pathogenic *PRKN* Variants detected in NIH-PD cohort by WGS.

In addition to the known pathogenic SNPs, we identified a novel intronic variant, c.7+5G>A, in a patient with EOPD, who also carried an E3-4 deletion (del) (Figure 1B (red font) and 1C). This variant was absent in MDSGene v3.5.95, gnomAD 2.1.1, and ClinVar ^6,29^. On further inspection, we determined that this variant lies in the predicted splice acceptor of intron 1 and would disrupt base pairing with the U1 snRNA (Figure 1C, left) ^30^. To phase the variants, we tested DNA from the proband’s unaffected sister and detected only the E3-4del, demonstrating that the proband’s variants are *in trans* (Figure 1C, right). Consistent with c.7+5G>A interfering with RNA splicing, a PCR product was not detected from the patient’s cDNA using primers that spanned the predicted splice site (Figure 1C, right). Based on these findings and additional functional studies described below, we considered the c.7+5G>A variant to be pathogenic. Altogether, 32 pathogenic variants were found in at total of 9 biallelic *PRKN* patients and 14 monoallelic *PRKN* patients by WGS.

### Screening for biallelic PRKN patients by genotyping array

We next assessed the accuracy of genotyping array for identifying *PRKN* biallelic patients, using the WGS genotyping as the gold standard. Genotyping array data (from the NeuroX or Neuro Consortium arrays ^23,24^) were available for 732/742 patients in the NIH cohort, including all but 1 patient with an identified *PRKN* mutation. Genotyping array identified 60% of pathogenic variants, including all *PRKN* deletions (12/12) and a third of pathogenic SNPs (6/18) (Figure 1D and Table 1). Notably, the missense variant p.R275W and deletions involving E2, E3, and/or E4 accounted for the majority (51.5%) of pathogenic *PRKN* mutations. All SNPs that were probed on the genotyping array, namely, p.R275W and p.R256C, were concordant with WGS calls. As expected, all SNPs without probes were missed. These included the frameshift mutations p.N34fs, p.N52fs, and p.113fs; a stop gain mutation, p.E409X; and the splice site mutation, c.7+5G>A. Deletion span was likewise concordant between genotyping array and WGS, as visualized in the Log R Ratio and B allele frequency plots of the genotyping array data with the WGS calls overlayed (Figure 1E). Altogether at least 1 pathogenic mutation was identified by genotyping array in 7/8 biallelic *PRKN* patients and 6/14 monoallelic *PRKN* patients. Thus, detection of at least 1 pathogenic *PRKN* mutation by NeuroX/NeuroChip genotyping arrays had a positive predictive value (PPV) of 54% and negative predictive value (NPV) of 99.9% (and a sensitivity of 87.5% and specificity of 99.2%) for identifying biallelic *PRKN* patients.

### Quantitative clinical data improves screening for biallelic PRKN patients

Next, we considered whether including quantitative clinical data with genotyping array data improves prediction of biallelic *PRKN* patients. To address this, we first determined which clinical data distinguished biallelic *PRKN* patients from monoallelic *PRKN* patients in our cohort. We reviewed charts for the biallelic and monoallelic *PRKN* groups identified by WGS, *LRRK2* p.G2019S (N=16) and *GBA* p.N409S (N=14) patient groups identified from genotyping array or WGS data, and the iPD patients with an available chart enrolled before each genetic PD patient. We also reviewed charts for a second group of 6 biallelic *PRKN* patients seen at the NIH (Supplementary Table 1).

We found that AAO was significantly younger and UPSIT was significantly higher for both biallelic *PRKN* groups compared to the iPD and monoallelic *PRKN* groups (Table 2), similar to what has been reported in other cohorts previously ^31,32^. By contrast, MoCA and UPDRS subscales were not significantly different among the groups (Table 2). Notably, all *PRKN* biallelic patients in our cohort had an AAO ≤ 38 and an UPSIT ≥ 20.

**Table 2:** Clinical characteristics of NIH-PD cohort.

| Clinical | iPD | GBA | LRRK2 | PRKN 1 mut | PRKN 2 mut<br>(group 1) | PRKN 2 mut<br>(group 2) | p-value<br>(unadjusted) | p-value<br>(corrected) |
| --- | --- | --- | --- | --- | --- | --- | --- | --- |
| AAO | 52.7±11.8 | 58.1±6.9 | 53.8±8.8 | 51.8±15.2 | 26.5±8.3*** | 30.7±5.5** | < 0.0001 | < 0.0001 |
| UPSIT | 20.3±7.9 | 20.7±8.2 | 23.5±7.1 | 20.8±6.7 | 31.9±6.5** | 33.7±4.5** | 0.0002 | 0.0014 |
| MoCA | 24.8±4.5 | 26.9±3.3 | 26.7±2.3 | 24.2±5.2 | 24.6±2.9 | 26.8±3.1 | 0.4733 | 3.3131 |
| UPDRSI | 2.9±2.9 | 3.8±1.8 | 3.3±2.3 | 3.9±3.0 | 3.0±3.0 | 1.8±1.7 | 0.7627 | 5.3389 |
| UPDRSII | 11.3±8.2 | 13.5±5.4 | 11.0±5.1 | 15.7±8.9 | 16.0±5.7 | 11.0±8.0 | 0.642 | 4.494 |
| UPDRSIII | 26.8±12.8 | 27.9±10.0 | 23.4±12.9 | 32.7±15.6 | 31.1±8.8 | 22.4±9.4 | 0.9547 | 6.6829 |
| UPDRSIV | 4.9±3.3 | 2.1±1.5 | 4.1±4.2 | 6.6±3.8 | 4.0±3.0 | 4.3±5.8 | 0.3235 | 2.2645 |

\*\* is < 0.01 and \*\*\* is <0.001 for Dunnett's multiple comparison test.
p-value (corrected) is a Bonferroni correction of the one-way ANOVA p-value.

UPSIT and AAO scores were separately available for a subset (334/739) of the WGS sequenced group. This allowed us to estimate how many patients with early onset and relatively preserved olfaction in our study have biallelic *PRKN* mutations. In total, 21 (6.3%) patients had AAO ≤ 38 and UPSIT ≥ 20. Of these, 3 (14.2%) were biallelic for *PRKN* mutations. Comparing the AAO and UPSIT for all available *PRKN* biallelic and monoallelic patients, biallelic *PRKN* patients formed a tight cluster, whereas monoallelic PRKN patients were distributed in a similar pattern as iPD patients (Figure 2A). Of note, 5 patients with variants that were novel or of uncertain significance (open circles in 2A) clustered with the other biallelic *PRKN* patients (solid circles in 2A), providing additional support for the variants’ pathogenicity.

**Figure 2.**
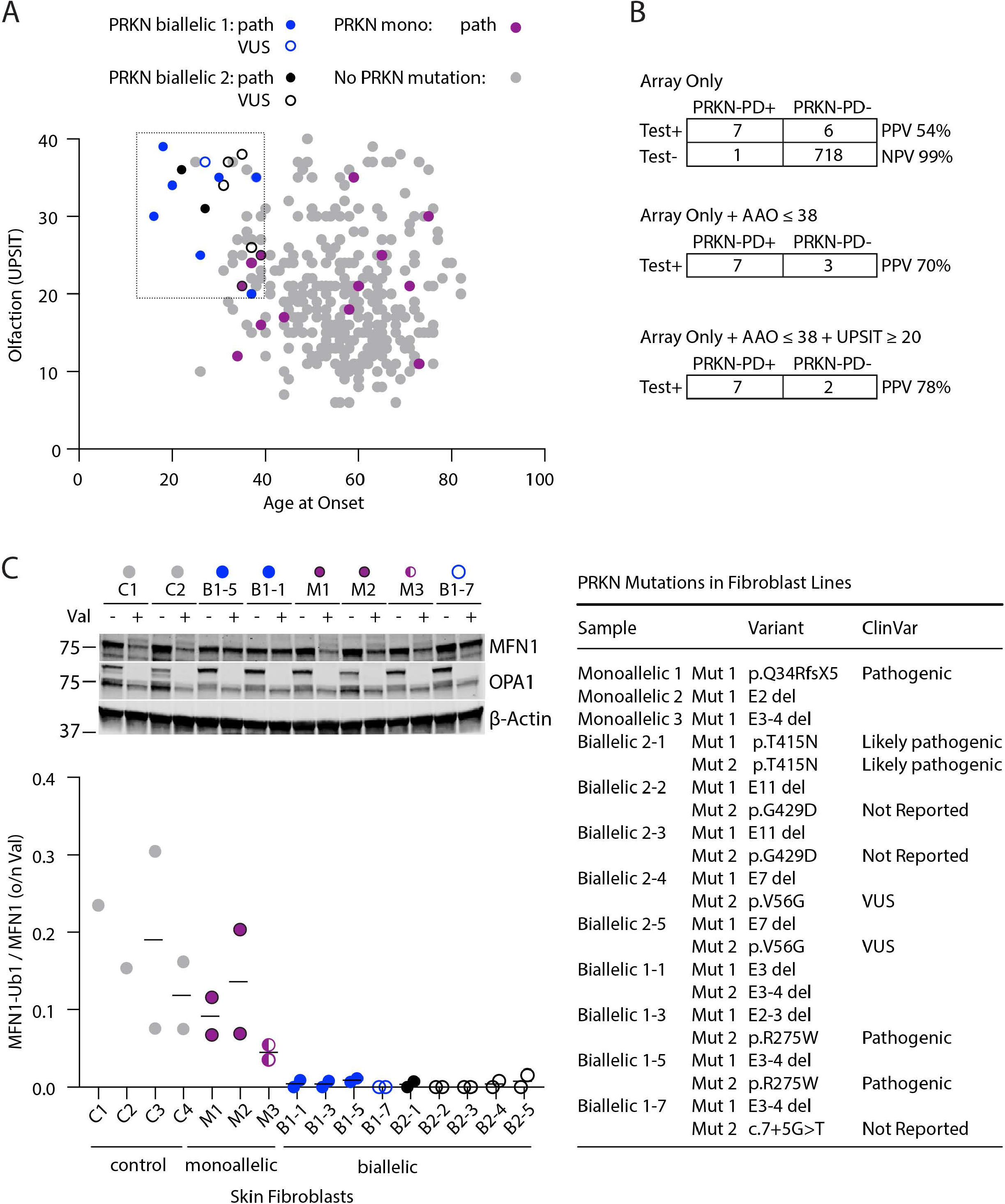
Biallelic and monoallelic *PRKN* mutation carriers can be distinguished by their phenotype and functional assays in their fibroblasts. (A) Scatterplot depicts scores from smell identification test (UPSIT, y-axis) and age at onset (x-axis) for genotyped PD patients in the NIH-PD cohort with no *PRKN* mutations (gray), 1 *PRKN* mutation (magenta), or 2 *PRKN* mutations (blue). A second group of patients with 2 *PRKN* mutations (black) was identified outside of the consecutive series. Those with a novel variant or variant of unclear significance (VUS) are shown as open symbols. 2 patients with 1 *PRKN* mutation and whose fibroblasts were evaluated in (C) have a black border. (B) Contingency tables show the positive and negative predictive values (PPV and NPV, respectively) to identify biallelic *PRKN* patients by detection of at least 1 *PRKN* variant by genotyping array with or without additional quantitative clinical information, using whole genome sequencing as the gold standard. (C) Patient fibroblasts described in table on right were treated with vehicle (DMSO) or 10 mM of valinomycin (val) overnight (o/n), separated on SDS-PAGE gels, and immunoblotted for the PARKIN substrate MFN1, as shown in representative blot (top). The ratio of MFN-Ub1 (corresponding to the slower migrating band) to MFN1 was calculated for each cell line treated with val. Each cell line was assayed in 2 biological replicates with exception of lines C1 and C2, which were assayed in 1 replicate each. All cell lines from *PRKN* carriers were from individuals with PD except for M3, who was the unaffected sibling of B1-7.

Together these data suggested that AAO and UPSIT scores can distinguish biallelic from monoallelic *PRKN* patients. Combining an AAO ≤ 38 threshold with genotyping array data increased the PPV for biallelic *PRKN* patients from 54% to 70% (Figure 2B). Adding an UPSIT ≥ 20 threshold further increased the PPV to 78%. Thus, by eliminating some single mutation carriers (false negatives), AAO and UPSIT cutoff values improved the predictive power of NeuroX/NeuroChip genotyping arrays for identifying biallelic *PRKN* patients.

### Assessing loss of PARKIN function in patient fibroblasts

3 patients in our study clustered phenotypically with biallelic *PRKN* patients but had only 1 detected *PRKN* mutation (Figure 2A, magenta data points within box). This raised the question of whether they may have a second undetected *PRKN* variant that was missed in our initial analysis of WGS data. Additionally, 5 patients had novel variants or variants annotated as of uncertain significance (open circles in 2A and C and Supplementary Figure 1), raising the question of whether the variants (namely, c.7+5G>A, p.V56G, and p.G429D) are pathogenic. To resolve definitively which patients had complete loss of *PRKN* function, we tested available fibroblast lines for ubiquitination of *PRKN* substrate Mitofusin-1 (MFN1) following *PRKN* activation by mitochondrial membrane depolarization ^12–16^. This assay has been used previously to differentiate biallelic *PRKN* carriers from healthy controls and *PRKN* carriers within a family, but to our knowledge has not been previously used to differentiate unrelated monoallelic and biallelic *PRKN* carriers ^16,20^. Altogether we tested fibroblast lines from 9 biallelic *PRKN* patients (including 5 with variants that were novel or of unclear significance), 1 patient with the novel c.7+5G>A *PRKN* mutation (*in trans* with an E3-4 deletion, as discussed above and in Fig. 1C), 3 monoallelic *PRKN* patients (including 2 that phenotypically clustered with biallelic PRKN patients (magenta with black border)), and 4 healthy controls (Figure 2C). Monitoring the change in MFN1-Ub1 / MFN1 ratio following depolarization with valinomycin distinguished all healthy controls and monoallelic *PRKN* patients from all biallelic *PRKN* patients (Figure 2C). Additionally, as a group, monoallelic *PRKN* carriers had an average MFN1-Ub1 / MFN1 ratio that was about half that of CTRLs, consistent with their possessing intermediate *PRKN* activity (Supplementary Figure 1A). Total MFN1 levels were also significantly decreased biallelic in *PRKN* cell lines compared to CTRL and monoallelic cell lines, consistent with the *PRKN*-dependent degradation of MFN1 (Supplementary Figure 1B). Two other measures of *PRKN* activity, levels of phospho-ubiquitin (Ser65) and mitophagy using the mt-Keima assay, failed to distinguish individual biallelic and monoallelic carriers, suggesting that these assays may not sensitive enough in fibroblasts with low endogenous *PRKN* levels (Supplementary Figure 1C -E).

In summary, functional assessment of patient fibroblasts resolved 3 VUSs (c.7+5G>A, p.V56G, and p.G429D) as pathogenic in 5 of our biallelic patients. Additionally, it ruled out a missed *PRKN* mutation in 2 early onset patients with 1 detected *PRKN* variant, validating our discovery cohort.

### Identifying *PRKN* copy number variants in the UK Biobank

We next used our high-confidence discovery cohort to develop sensitive screening algorithms for identifying *PRKN* CNVs in genotyping array data (described in the methods). These optimized algorithms were then applied to genotyping array data from 488,264 subjects in the UK Biobank. Collectively, they flagged 25,906 subjects in the UK Biobank as potentially carrying a CNV, 2,687 of which were confirmed to have a CNV on visual inspection of the Log R Ratio and B allele frequency plots. From the whole cohort a deletion was detected in 0.30% of samples, and a duplication in 0.25% of samples. This was close to the percentage of deletions (0.34%) and duplications (0.26%) that we predicted based on visual inspection of every 100^th^ sample. Altogether, we estimated that ∼92% of detectable CNVs were identified in the UK biobank.

To visualize the CNVs, we generated heatmaps for all single deletions and duplications, as well as the average values for each class of CNV (Figure 3). E2del and E2dup were the most common class of deletion and duplication, respectively, together accounting for 48.9% of all CNVs. Interestingly, while the lengths of E2 dels were highly variable, the lengths of E2dups were more uniform. Although genotyping array does not provide the resolution to define exact breakpoints, this suggests that E2 dups may be composed of fewer distinct CNVs compared to E2dels. To provide additional support for this observation, we examined *PRKN* E2dups and E2dels called from WGS data in the gnomAD SVs v2.1 database ^33^. Consistent with results from the UK Biobank, the most frequent E2dup in the European population accounted for 40% of all E2dups, whereas the most frequent E2del accounted for only 13.4% of E2dels. This confirmed that there is greater diversity among E2dels than E2dups (Figure 3).

**Figure 3.**
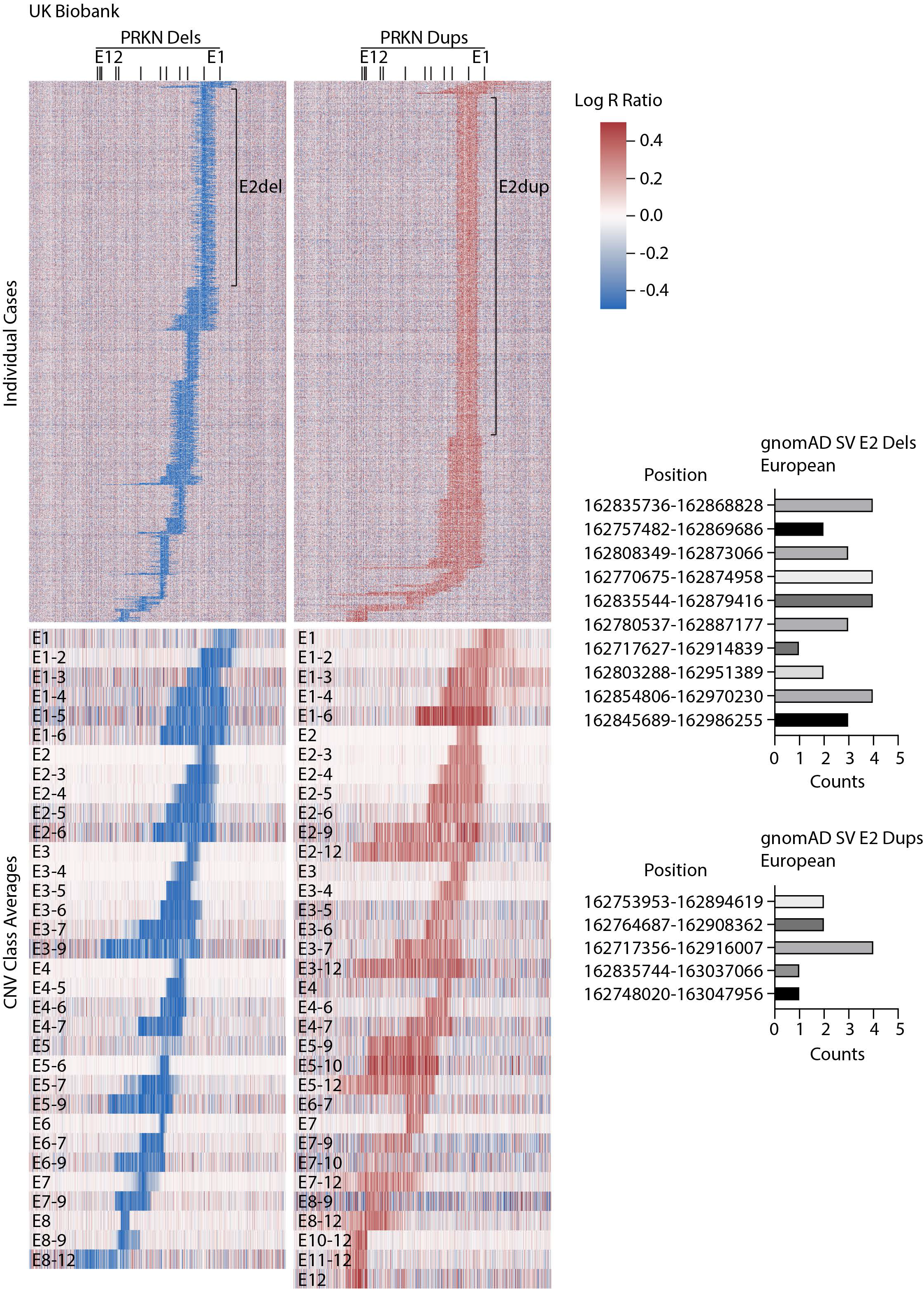
Identification of *PRKN* CNVs in UK Biobank from genotyping array data. Heat maps represent Log R Ratio across the *PRKN* locus (on the x axis). Deletions (dels) are shown on the left and duplications (dups) on the right. The heat maps of the top depict all individual cases (on the y-axis) in which a single CNV was detected. The locations of E2dels and E2dups are shown. Heat maps on bottom represent the average Log R Ratios for each class of CNV. Graphs on right show the distributions of E2dels (top) and E2dups (bottom) for the non-Finnish European population in the gnomAD SV database.

We next compared the distribution of the 2,693 CNVs (identified in 2,687 carriers) in the UK Biobank to 3 other studies/databases, each of which has more than 100 CNVs: the deCode study of the Icelandic population, which detected CNVs from genotyping array (993 CNVs) ^9^; the MDS Gene database (435 CNVs), a curated database of the published literature ^6^; and the gnomAD SVs v2.1 database ^33^, which called CNVs from WGS data (171 CNVs) (Figure 4A and B and Supplementary Table 2). In all studies, most deletions and duplications involved E2, E3, or E4 (88.2% in UK Biobank, 96.4% in deCode, 65.7% in MDS gene, and 94.7% in gnomAD). Notably, the 2 most common classes of deletions in UK Biobank, E2del and E3-4del, were among the top 4 in MDSGene, the top 3 in deCode, and the top 3 in gnomAD. An E6-9del that was particularly common in deCode (21% of deletions) was uncommon in UK Biobank (1.2% of deletions) and absent in MDS Gene and gnomAD. This suggests that the high E6-9del frequency may be due to a founder effect in the Icelandic population.

**Figure 4.**
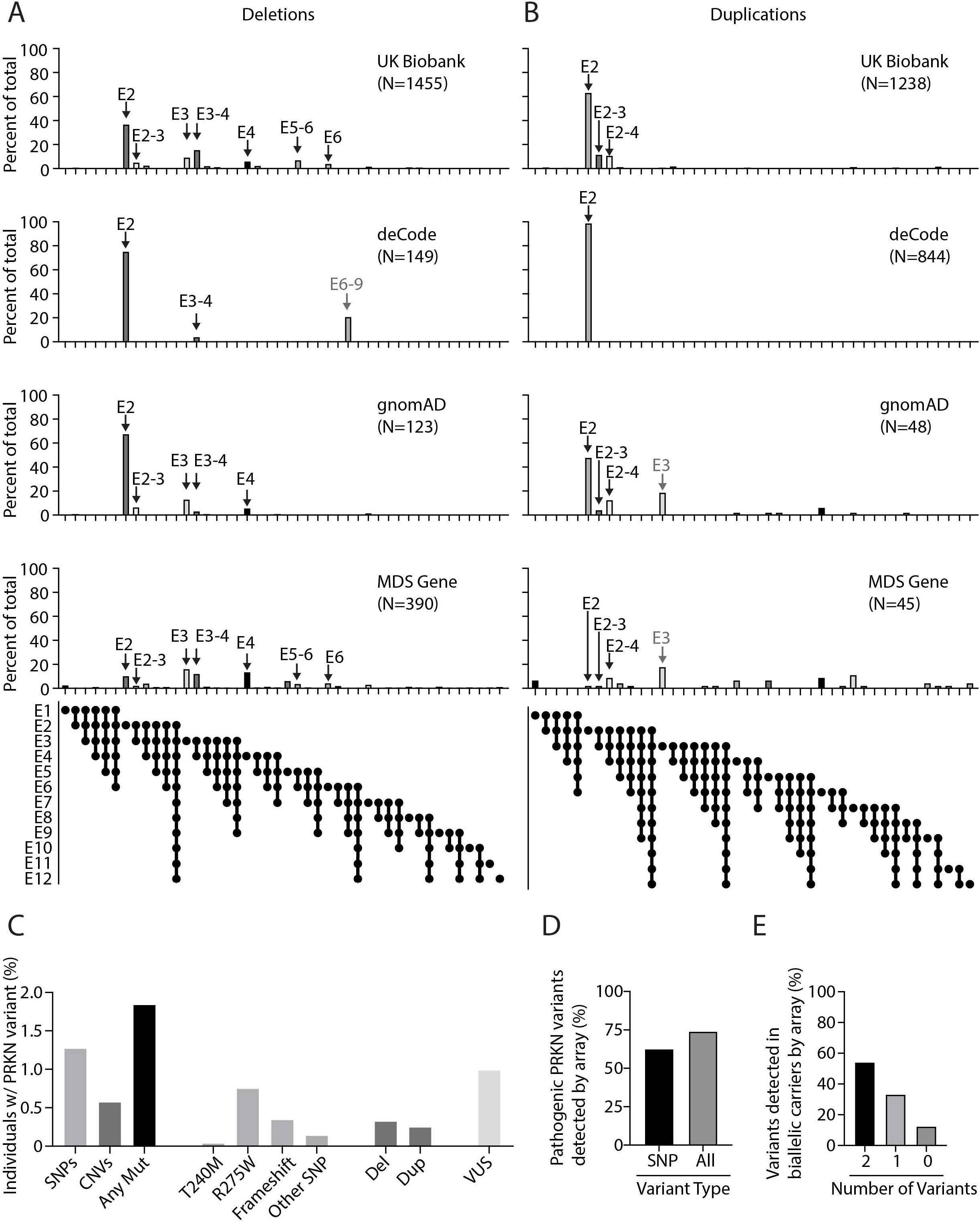
Distribution of *PRKN* CNVs in UK Biobank and other studies. (A and B) Graphs depict distributions of *PRKN* deletions (A) and duplications (B) in the UK Biobank detected in this study vs. those reported in the deCode study, the gnomAD SV database, and the MDS gene database. Major peaks identified in the UK Biobank are indicated by black arrows. Gray arrows signify major peaks (>15% of dels or dups, respectively) that were detected in one of the other databases and were rare in the UK Biobank (<1%). (C) Graph represents *PRKN* variants detected in the UK Biobank for participants with both WES and genotyping array data. (D) Graph represents the number of pathogenic *PRKN* variants detected by genotyping array for participants with both WES and genotyping array data. (E) Graph represents the number of pathogenic *PRKN* variants detected by genotyping array in biallelic *PRKN* variant carriers with both WES and genotyping array data.

Among duplications, the E2dup was the most common duplication in the UK Biobank, the deCode study, and gnomAD, although it was rare in the MDS Gene database. Overall, duplications were less common in the literature as represented in the MDS Gene Database than in the population-based studies, possibly reflecting a bias in their detection or reporting (10% of CNVs in MDS Gene Database vs. 28% in gnomAD, 48% in the UK Biobank, and 70% in deCode). Interestingly, an E3 duplication accounted for 18.8% of duplications in gnomAD and 17.8% of duplications in MDS gene but was absent in the UK Biobank. In gnomAD all 9 E3 duplications had the same breakpoints (6:162638588-162730211) and the allelic frequency was highest for the African/African-American population, suggesting that they may reflect a founder effect.

Overall, the distribution in CNVs was similar in the UK Biobank as in other studies and databases with *PRKN* CNV data with most CNVs affecting exons 2, 3, or 4. Founder effects may account for the increased prevalence of E2dups in the UK and Icelandic populations relative to others. Similarly, an E6-9 del and an E3 dup may be due to founder effects in the Icelandic and African-American/African populations, respectively.

### Estimating pathogenic *PRKN* variant frequency in the UK Biobank

To obtain a more complete view of the prevalence of pathogenic *PRKN* variants in the UK population, we next examined the frequency of pathogenic *PRKN* missense variants in the UK Biobank. The UK Biobank genotyping array contains probes for 4 known pathogenic *PRKN* missense variants: p.R275W, p.C253Y, p.T240M, and p.K211N. To validate these probes, we benchmarked their performance against WES data from 192,490 participants genotyped on both platforms. Genotyping array calls agreed with WES calls for p.R275W and p.T240M variants in most samples (98.5% and 100%, respectively), but were less reliable for p.C253Y and p.K211N variants (4.4% and 70%, respectively). Thus, we restricted further analysis of genotyping array data to p.R275W and p.T240M variants.

Assessed in the entire UK Biobank from genotyping array data, p.R275W and p.T240M mutations had allelic frequencies of 0.0039 and 0.00022, respectively. These were similar to their allelic frequencies in the subset of samples with WES data (0.0038 and 0.00019, respectively), as well as their frequency in the non-Finnish European population in gnomAD v2.1.1 database (0.0033 and 0.00023, respectively). In the UK Biobank, p.R275W was more common in Europeans, whereas p.T240M was more common in non-Europeans. The latter finding may reflect the higher allelic frequency of p.T240M in the South Asian population, as also seen in gnomAD v2.1.1 (0.0017 vs. 0.00023).

As genotyping array probes reliably for only two pathogenic mutations, we also estimated the prevalence of pathogenic or loss of function (LOF) *PRKN* variants in samples with WES data. Altogether, 37 pathogenic or LOF *PRKN* SNPs were identified in 1.27% of samples (Figure 4C and Supplementary Figure 3). The most common pathogenic variant was p.R275W, which was present in 0.75% of samples. 217 rare *PRKN* missense variants that were annotated as VUS or not annotated were present in an additional 1% of samples (Supplementary Table 4). Combining WES + genotyping array estimates, we found a pathogenic *PRKN* mutation in 1.84% of samples. 24 (∼1/8400) carried biallelic pathogenic variants. Genotyping array captured most of these variants (74.1%) and identified at least 1 variant in most biallelic carriers (87.5%) (Figure 4D and E).

### Risk of PD in single PD mutation carriers

Altogether, we detected single pathogenic mutations in 6,612 of samples by genotyping array, more than what has been reported in any previous study or pooled studies subjected to meta-analysis ^10^. Given its size, we reasoned it may be particularly valuable for testing whether heterozygous *PRKN* variants increase the risk of developing PD. We also tested whether *PRKN* carriers were more likely to have a parent with PD, given one of the parents is an obligate *PRKN* carrier in most cases. Additionally, while the median age of UK Biobank participants at enrollment (i.e., 58) is less than the average age of PD onset, their parents age is likely greater than the average age of PD onset. Thus, PD prevalence is likely higher in the parents than the probands.

Among participants with 0 or 1 detected heterozygous *PRKN* mutation, a total of 3,465 reported having PD, and 17,675 reported having a parent with PD (Table 3). Those carrying 1 detected heterozygous *PRKN* mutation were as likely as those without a mutation to have PD (OR = 1.18, 95%CI = 0.88 – 1.54, p-value = 0.24). Likewise, those with 1 mutation were as likely as those without mutation to have a parent with PD (OR = 1.06, 95%CI = 0.93 – 1.20, p-value = 0.39). We obtained similar findings testing each class of mutation individually.

**Table 3:** Odds of PD or parent with PD in *PRKN* heterozygous mutation carriers in UK Biobank.

| Mutation | Mut_Cases | Mut_CTRLs | NoMut_Cases | NoMut_CTRLs | Odds | CI | p-value |
| --- | --- | --- | --- | --- | --- | --- | --- |
| T240M | 3 | 208 | 3455 | 484125 | 2.02 | 0.41-5.99 | 0.19 |
| R275W | 34 | 3705 | 3389 | 476062 | 1.29 | 0.89-1.81 | 0.14 |
| AnySNP | 37 | 3913 | 3428 | 480963 | 1.33 | 0.93-1.84 | 0.10 |
| DEL | 11 | 1422 | 3454 | 483454 | 1.08 | 0.54-1.95 | 0.75 |
| DUP | 7 | 1222 | 3458 | 483654 | 0.80 | 0.32-1.66 | 0.73 |
| AnyCNV | 18 | 2644 | 3447 | 482232 | 0.95 | 0.56-1.51 | 1.00 |
| **AnyMut** | **55** | **6557** | **3410** | **478319** | **1.18** | **0.88-1.54** | **0.24** |

Odds of parent with PD in UK Biobank Participants with Microarray (N = 488,341)
| Mutation | Mut_Cases | Mut_CTRLs | NoMut_Cases | NoMut_CTRLs | Odds | CI | p-value |
| --- | --- | --- | --- | --- | --- | --- | --- |
| T240M | 9 | 202 | 17644 | 469936 | 1.19 | 0.53-2.30 | 0.58 |
| R275W | 135 | 3604 | 17363 | 462088 | 1.00 | 0.83-1.18 | 1.00 |
| AnySNP | 144 | 3806 | 17531 | 466860 | 1.01 | 0.85-1.19 | 0.93 |
| DEL | 65 | 1368 | 17610 | 469298 | 1.27 | 0.97-1.63 | 0.07 |
| DUP | 43 | 1186 | 17632 | 469480 | 0.97 | 0.69-1.31 | 0.88 |
| AnyCNV | 108 | 2554 | 17567 | 468112 | 1.13 | 0.92-1.37 | 0.23 |
| **AnyMut** | **252** | **6360** | **17423** | **464306** | **1.06** | **0.93-1.20** | **0.39** |

Odds of PD in UK Biobank Participants with WES and Microarray (N = 200,606)
| Mutation | Mut_Cases | Mut_CTRLs | NoMut_Cases | NoMut_CTRLs | Odds | CI | p-value |
| --- | --- | --- | --- | --- | --- | --- | --- |
| Frameshift_SNP | 2 | 683 | 1372 | 198549 | 0.42 | 0.05-1.54 | 0.35 |
| AnySNP | 15 | 2517 | 1359 | 196715 | 0.86 | 0.48-1.43 | 0.71 |
| **AnyMut** | **23** | **3648** | **1351** | **195584** | **0.91** | **0.58-1.38** | **0.76** |

Odds of parent with PD in UK Biobank Participants with WES and Microarray (N = 200,606)
| Mutation | Mut_Cases | Mut_CTRLs | NoMut_Cases | NoMut_CTRLs | Odds | CI | p-value |
| --- | --- | --- | --- | --- | --- | --- | --- |
| Frameshift_SNP | 29 | 656 | 7465 | 192456 | 1.14 | 0.76-1.65 | 0.48 |
| AnySNP | 104 | 2428 | 7390 | 190684 | 1.11 | 0.90-1.35 | 0.32 |
| **AnyMut** | **152** | **3519** | **7342** | **189593** | **1.12** | **0.94-1.31** | **0.19** |

As genotyping array data missed an estimated 27% of *PRKN* mutations in the UK Biobank, we also assessed the subset of 200,606 participants with WES + genotyping array data and 0 or 1 *PRKN* pathogenic *PRKN* variants. Although this was only about half of the total UK Biobank, it still represented more PRKN carriers than previously analyzed and had the advantage of near complete genotyping of *PRKN* SNPs and CNVs. Consistent with the genotyping array only dataset, participants with 1 detected *PRKN* mutation by WES + genotyping array were as likely as those with no mutation to have PD (OR = 0.91, 95%CI = 0.58 – 1.38, p-value = 0.76) or a parent with PD (OR = 1.12, 95%CI = 0.94 – 1.31, p-value = 0.19). Together our findings show that monoallelic heterozygous *PRKN* mutations do not significantly increase the risk of having PD or a parent with PD.

## DISCUSSION

In this study, we evaluated the effectiveness of genotyping array to identify *PRKN* mutations in a large PD cohort and a population scale cohort. In both cohorts, we found that genotyping array identified most pathogenic *PRKN* variants and detected at least 1 pathogenic variant in 87.5% of biallelic carriers. Additionally, most patients in which at least 1 pathogenic *PRKN* mutation was detected by genotyping array were biallelic carriers. Inclusion of AAO and UPSIT further improved the accuracy of genotyping array screening by eliminating false positives. Finally, sampling nearly half million individuals in the UK Biobank, we also found that *PRKN* mutations were common in the general population, and that carrying a single heterozygous *PRKN* mutation did not significantly increase risk of having PD or a parent with PD.

Notably, genotyping array detected the majority of *PRKN* mutations in both the NIH PD cohort and the UK Biobank. In the NIH PD cohort, genotyping array identified all deletions that were also called by WGS, as well as a third of *PRKN* SNPs. Genotyping array performed similarly in the UK Biobank, identifying an estimated 74.1% of pathogenic variants, including most pathogenic SNPs. Notably, similar results were achieved in these two cohorts even though different genotyping platforms were used (Illumina in NIH-PD vs. Affymetrix in the UK Biobank).

Genotyping array performed well compared to next-generation sequencing, identifying at least 1 *PRKN* mutation in 87.5% of *PRKN*-PD patients. Likewise, if 1 mutation was identified, most patients in the NIH-PD cohort had a second pathogenic *PRKN* mutation. This probability was further increased if age at onset was ≤ 38 or olfaction was relatively preserved (UPSIT ≥ 20). Thus, even though genotyping array did not provide complete genotyping of *PRKN*, it served as an effective screening tool for identifying *PRKN*-PD patients. Together these results suggest a *PRKN*-PD screening framework in which genotyping array + PD status + AAO could be used in initial screening, followed by secondary screening with *PRKN* sequencing, and (optionally) functional testing of skin fibroblasts to resolve *PRKN* variants of unclear significance. Together this framework could efficiently and confidently identify *PRKN*-PD patients for targeted trials, such as those aimed at gene or cell replacement.

The strong performance of genotyping arrays in our cohorts was due to good coverage of the *PRKN* locus by the arrays used and the high frequency of the missense variant p.R275W and CNVs involving E2, E3, and/or E4 in our cohorts. Together these accounted for 51.5% of mutations in the NIH PD cohort and ∼72% of mutations in the UK Biobank. Similar results are likely achievable with other genotyping arrays in populations with high European ancestry, provided they genotype the p.R275W variant and they have adequate coverage of E2, E3, and E4 to determine their copy number (typically at least 4 probes per exon). For arrays that currently do not have this coverage, the addition of ∼13 probes would allow for screening of most *PRKN*-PD patients in populations of predominantly European-ancestry for *PRKN*-PD targeted trials.

In non-European populations, the p.R275W variant is less frequent, and, therefore, genotyping arrays may not capture the majority of *PRKN* variants. This highlights the need for *PRKN* sequencing in populations with non-European ancestry, and inclusion of *PRKN* SNPs from these populations on genotyping arrays. The novel NeuroBooster array and GP2, the Global Parkinson’s Genetics Program, will help address these needs ^34^. Nonetheless, genotyping arrays are still likely to be effective for identifying a substantial portion of *PRKN* CNVs in non-European populations. We found CNVs were present in > 0.5% of participants in the UK Biobank, regardless of ancestry. This likely reflects the diversity of CNVs detected. The most common classes of deletions, for instance, were composed of several CNVs of distinct length, suggesting that they were generated by several independent recombination events. This diversity makes it more likely that CNVs are evenly distributed among populations. The frequency and diversity of *PRKN* CNVs is likely related the large size of the gene (1.38 Mb), making recombination events more likely.

This was the first population scale study to estimate the frequency of pathogenic *PRKN* mutations using methods that capture both most SNPs (by WES) and most CNVs (by genotyping array). Altogether we detected a pathogenic *PRKN* mutation in 1.8% of participants in the UK Biobank. Although higher than some estimates based on SNP frequency alone, our estimate is generally in line with those from smaller studies that have employed methods to capture most SNPs and CNVs. Yu et al., for instance, recently found that 1.8% of control subjects carried a *PRKN* mutation ^35^. Along these lines, we found biallelic *PRKN* mutations in ∼1/8400 UK Biobank participants, which would correspond to ∼7,900 biallelic *PRKN* carriers in the UK and close to a million world-wide. Together these findings suggest that pathogenic *PRKN* variants are common in the general population.

Strikingly, the clinical significance of many *PRKN* variants remains unknown. Although individually rare, 217 *PRKN* missense variants of unclear significance or without annotation in ClinVar were found in 1% of UK Biobank samples. This represents about a 1/3 of variants that affect an exon and are not annotated as benign. In the NIH-PD cohort we found that assaying *PRKN* function in *PRKN* fibroblasts can differentiate unrelated samples with 1 or 2 heterozygous pathogenic mutations. This allowed us to resolve the pathogenicity of 3 variants that were novel or annotated as VUS. This also allowed us to rule out cryptic pathogenic variants in 2 patients with single pathogenic *PRKN* variants and a *PRKN*-PD phenotype (i.e., early AAO and preserved olfaction). Functional studies may help to clarify which of the 217 missense VUSs in the UK Biobank are loss of function and therefore likely pathogenic.

Whether heterozygous *PRKN* variants increase the risk of PD has been unclear. A large association study of the Icelandic population involving 105,749 genotyping array samples found that heterozygous *PRKN* CNVs increased the odds of PD (with an Odds Ratio of 1.69) ^9^. A recent study nominally replicated this association in a case-control study and meta-analysis but found that the association is lost if missed mutations in biallelic *PRKN* carriers are taken into account ^10^. Here, using the largest dataset to date (a subset of which had near complete coverage of known pathogenic *PRKN* variants), we did not find an association between heterozygous *PRKN* mutations and PD risk. Similarly, heterozygous *PRKN* mutations did not increase the odds of having a parent with PD. This is notable as 1 parent of most *PRKN* carriers is an obligate carrier, and most parents are beyond the age of onset for most PD cases. The number of parents with PD in the UK Biobank is much larger than the number of PD cases (17,675 vs. 3,465), which increases the power to detect an association. In contrast to the lack of association between heterozygous *PRKN* variants and PD in the UK Biobank, association of common variants with PD risk has been replicated in the UK Biobank previously ^36^.

In summary, through comprehensive analysis *PRKN* mutations in a large PD cohort and a population scale cohort, we validated genotyping array screening for the detection of biallelic *PRKN* patients. Additionally, we demonstrated that functional assays in patient cell can resolve *PRKN* VUSs in biallelic patients and can rule out second cryptic variants in patients with 1 heterozygous pathogenic mutation. Finally, we demonstrate that heterozygous *PRKN* variants are common in the population but do not increase the risk of PD.

## Study Limitations

We detected ∼92% of CNVs in the UK Biobank that can be identified by visual inspection. However, small CNVs that were not well covered by probes on the genotyping array may have been missed, particularly those involving single exons 1, 5, 9, 10, and 12. These likely make up a small proportion of *PRKN* CNVs, however, as they account for only 0.6% of all CNVs in gnomAD (all are absent except E5 dups) (Supplementary Table 2)). These would tend to underestimate the prevalence of *PRKN* mutations in the UK population but are not predicted to affect the association between single pathogenic *PRKN* variants and PD risk, as they are as likely to occur in cases as controls.

## ACKNOWLEDGEMENT

This research has been conducted using the UK Biobank Resource under Application Number 33601. This work utilized the computational resources of the NIH HPC Biowulf cluster (http://hpc.nih.gov).

## SUPPLEMENTARY MATERIAL

**Supplementary Figure 1. Assessing PARKIN function in patient fibroblasts**. (A and B) Graphs depict additional data quantified from representative blots in Figure 2C. The ratio of MFN1-Ub1 to MFN1 levels were quantified at the group level for controls (CTRLs), carriers of 1 *PRKN* mutation, and patients with 2 *PRKN* mutations (characteristics of cell lines are shown in Figure 2C). (B) MFN1 levels relative to DMSO treated samples for groups in (A). ns, non-significant; ^**^, p-value < 0.01; ^***^, p-value < 0.001; ^****^, p-value < 0.0001. (C) A subset of control (C2 and C5), *PRKN* 1 mutation (M2 and M3), or *PRKN* 2 mutations (B1-7 and B2-5) cell lines were transduced with the mt-Keima mitophagy reporter and treated with vehicle or 10 mM valinomycin (val) overnight. Cells were analyzed by flow cytometry. mt-Keima is a fluorescent protein that is differentially excited depending on the surrounding pH. The ratio of pH8/pH4 indicates the proportion of fluorescent protein in free mitochondria vs. mito-lysosomes as a measure of mitophagic flux. Experiments were performed in biological duplicate. (D) Lysates from experiment described in Figure 2 were immunoblotted for the PINK1 substrate pS65-Ub. PARKIN amplifies production of this substrate in a feed-forward cycle with PINK1. Each cell line was analyzed in a single replicate. Assays in (C) and (D-E) did not allow individual cell lines to be differentiated based on the number of pathogenic *PRKN* variants.

**Supplementary Table 1: Characteristics of all biallelic *PRKN* patients evaluated at NIH**.

**Supplementary Table 2: Proportion of copy number variants (CNVs) in UK Biobank (analyzed in this study), the deCode study, MDS gene database, and the gnomAD SV v2.1 database**.

**Supplementary Table 3: Frequency of pathogenic *PRKN* variants in UK Biobank samples with WES and genotyping array**.

**Supplementary Table 4: Missense variants in UK Biobank without annotation or annotated as VUS**.

