## Supplementary figures and images for "Heterozygous *PRKN* mutations are common but do not increase the risk of Parkinson’s disease"

### Supplementary Figure 1

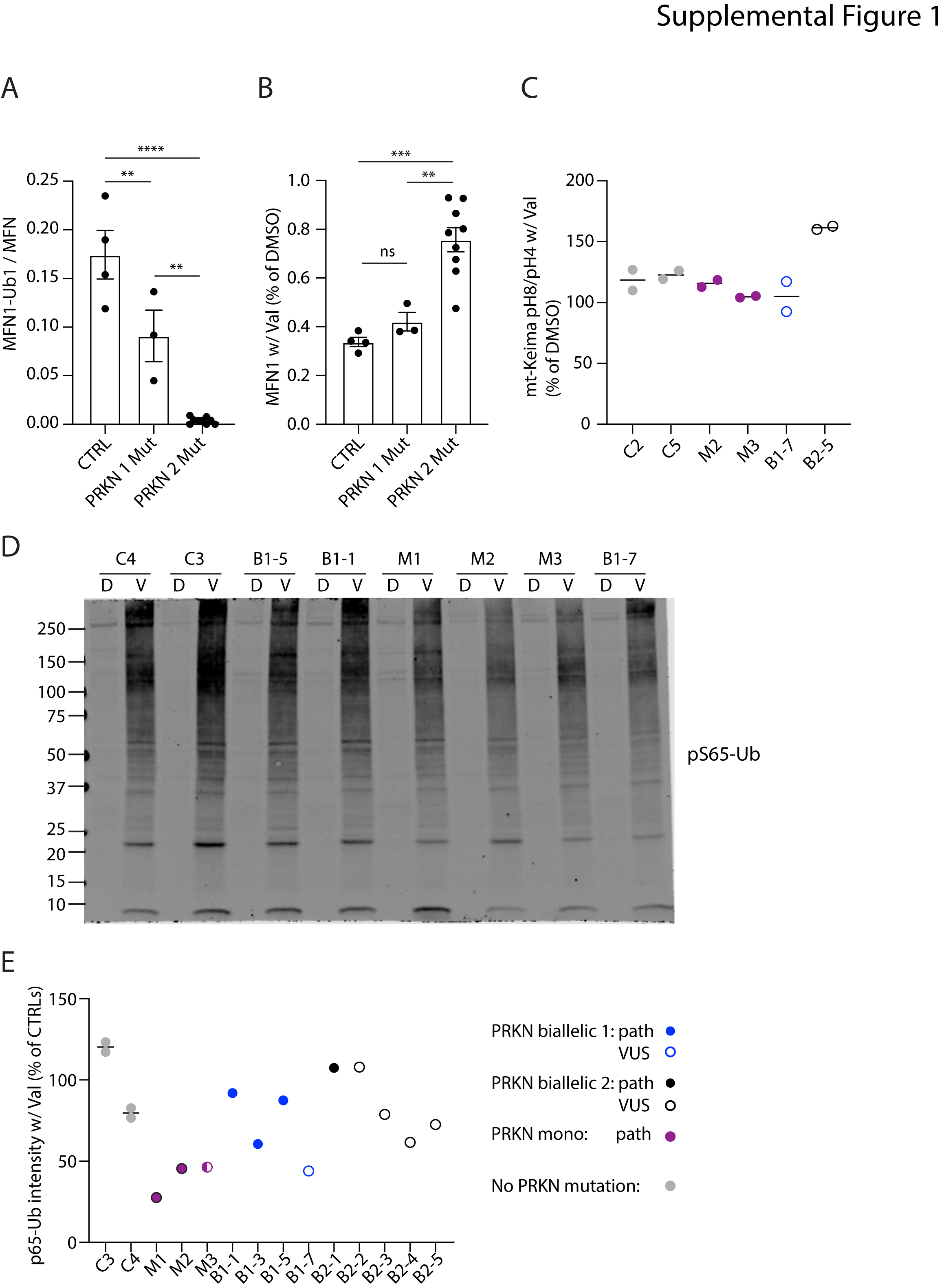
